# Sudden rise in COVID-19 case fatality among young and middle-aged adults in the south of Brazil after identification of the novel B.1.1.28.1 (P.1) SARS-CoV-2 strain: analysis of data from the state of Parana

**DOI:** 10.1101/2021.03.24.21254046

**Authors:** Maria Helena Santos de Oliveira, Giuseppe Lippi, Brandon Michael Henry

**Affiliations:** Department of Statistics, Federal University of Parana, Curitiba, Brazil; Section of Clinical Biochemistry, University of Verona, Verona, Italy; The Heart Institute, Cincinnati Children’s Hospital Medical Center, Cincinnati, OH, USA

**Author notes:** Funding: None. Conflicts of Interests: None. **Correspondence to**: Brandon Michael Henry, MD, Cardiac Intensive Care Unit, The Heart Institute, Cincinnati Children’s Hospital Medical Center, 3333 Burnet Ave., Cincinnati, OH 45229, USA.

**Keywords:** case fatality rate, variant of concern, Brazil, epidemiology, COVID-19

## Abstract

Brazil is currently suffering a deadly surge of severe acute respiratory syndrome coronavirus 2 (SARS-CoV-2) infections, which has been attributed to the spread of a new strain known as P.1 (B.1.1.28.1). In this investigation, we analyzed coronavirus disease 2019 (COVID-19) public health data from Parana, the largest state in southern half of Brazil, between September 1, 2020 and March 17, 2021, to evaluate recent trends in case fatality rates in different age groups. A total of 553,518 cases of SARS-CoV-2, 8,853 currently registered as fatal, were finally included in our analysis. All age groups showed either decline or stabilization of the case fatality rates (CFRs) between September 2020 and January 2021. In February 2021, an increase in CFR for almost all age groups could be instead observed. All groups above 20 years of age showed statistically significant increases in CFR when diagnosed in February 2021 as opposed to January 2021. Patients aged 20-29 years experienced a tripling of their CFR, from 0.04% to 0.13%, while those aged 30-39, 40-49, 50-59 experienced approximate CFR doubling. Individuals between 20 and 29 years of age whose diagnosis was made in February 2021 had an over 3-fold higher risk of death compared to those diagnosed in January 2021 (Risk Ratio (RR): 3.15 [95%CI: 1.52-6.53], p<0.01), while those aged 30-39, 40-49, 50-59 years experienced 93% (1.93 [95%CI:1.31-2.85], p<0.01), 110% (RR: 2.10 [95%CI:1.62-2.72], p<0.01), and 80% (RR: 1.80 [95%CI:1.50-2.16], p<0.01) increases in risk of death, respectively. Notably, the observed CFR increase coincided with the second consecutive month of declining number of diagnosed SARS-CoV-2 cases. Taken together, these preliminary findings suggest significant increases in CFR in young and middle-aged adults after identification of a novel SARS-CoV-2 strain circulating in Brazil, and this should raise public health alarms, including the need for more aggressive local and regional public health interventions and faster vaccination.

## INTRODUCTION

Brazil is in the midst of a deadly surge of severe acute respiratory syndrome coronavirus 2 (SARS-CoV-2) infections, with daily deaths recently peaking at 2,841 on March 16, 2020.^1^ To-date, there have been over 11 million cases in Brazil, resulting in over 290,000 deaths.^1^ In January 2021, a novel strain of SARS-CoV-2, called P.1 (formerly known as B.1.1.28.1), was first identified in Japan among returning travelers from Amazonas state in the north of Brazil, and has rapidly become ubiquitous throughout the country.^2^ Compared to its ancestral strain B.1.1.28, P.1 acquired several mutations in the spike protein, including E484K and N501Y, which would make it biologically similar to the B.1.351 strain identified in South Africa.^2^ The presence of the N501Y mutation would suggest that this variant may also be characterized by ∼50% increased transmissibility^3^, with current data suggesting P.1 to be associated with 1.4–2.2 higher transmissibility.^4^ The E484K has been reported to be associated with humoral immune escape, as observed in reports of decreased neutralization efficacy of convalescent plasma and sera from vaccinated individuals, as well as reports of re-infection.^5^ In particular, the P.1 strain has been shown to escape neutralization by vaccine-induced humoral immunity in pseudoviral models.^6,7^

Given the mutations acquired by P.1, questions have emerged that in addition to enhanced transmissibility and potential immune escape, it may be associated with enhanced virulence and pathogenicity. Indeed, it is reasonable to speculate that enhanced binding of the mutated virus to its host receptor (angiotensin converting enzyme 2 (ACE2)), may result in increased viral load, with resultant increased virulence.^8^ To-date, limited evidence is available on clinical outcomes of P.1. Nonetheless, media reports from Brazil have suggested that in the current surge, increased morbidity and mortality is being observed among younger individuals.^9^ However, earlier in the pandemic, we had previously reported higher mortality rates in younger individuals (<60 years old) in Brazil as compared to China and Italy.^10^

In this investigation, we probed epidemiologic data from the state of Parana, in the south of Brazil, where the P.1 variant was first officially identified on February 16^th^, 2021^11^, to assess any recent trends in mortality data among different age-grouped populations. As of March 3^rd^, 2021, surveillance data had indicated that P.1 accounted for 70.4% of tested samples that were screened for the new variant in Parana.^12^ We choose a single state to reduce the number of confounders and heterogeneity between geographical regions in Brazil, but enabling analysis of a large enough sample to garner sufficient data for an informative analysis. With over 11 million habitants, Parana has the largest population out of the states in the southern region of Brazil, and the fifth largest in the entire country. The first cases of COVID-19 in the state were confirmed on March 12th, 2020, and one year later, Parana has confirmed over 780,000 cases of SARS-CoV-2 infection.

## METHODS

Age, date of diagnosis, and current status data on individuals with confirmed SARS-CoV-2 infection data was obtained from the Parana State Secretary for Health for a period from September 1^st^, 2020 to March 17^th^, 2021. Data was organized by age groups and month of diagnosis, and monthly case fatality rate (CFR) with 95% confidence intervals (95%CI) was generated. Additionally, risk ratios (RRs) with 95%CI were calculated to quantify the difference in mortality between the months of January and February 2021. Monthly CFR was defined as the proportion of individuals diagnosed in a particular month who were registered as deceased by March 17th, 2021. Statistical analysis was performed using the R software for statistical computing (version 6.3.2) and the package epiR: Tool for the Analysis of Epidemiological Data (version 1.0–14). The study was carried out in accordance with the Declaration of Helsinki, under the terms of relevant local legislation.

## RESULTS

The obtained dataset contained data regarding 553,518 individuals infected with SARS-CoV-2 in Parana between September 2020 and March 2021, 8,853 registered as deceased. Summary of results presented as CFR by age group over the study period is presented in Figure 1. All age groups showed either decline or stabilization of CFRs between September 2020 and January 2021 (Figure 1). The month of February 2021 shows a contrasting increase in fatality among almost all age groups. All groups above 20 years of age showed statistically significant increases in CFR when diagnosed in February 2021 as opposed to January 2021 (Table 1). Patients aged 20-29 years experienced a tripling of their CFR from 0.04% in January 2021 to 0.13% in February 2021. Patients aged 40-49 years experienced a doubling in CFR between January 2021 and February 2021, increasing from 0.43% to 0.90%. Similarly, patients aged 30-39 years and 50-59 years, experienced a near doubling of their CFRs between January 2021 and February 2021, while older patients, aged >60 years, experienced significant increases, albeit at a less dramatic rate of change (Table 1).

**Table 1.** Case fatality rates and estimated death risk ratios for SARS-CoV-2 infected persons in Parana, Brazil between January 2021 and February 2021.

| **Age Group**<br>**(years)** | **Case Fatality Rate (95% CI)** |  | **Death Risk Ratio (95%**<br>**CI)** | **p-value** |
| --- | --- | --- | --- | --- |
|  | **January 2021** | **February 2021** |  |  |
| 0-5 | 0.10% | 0.12% | 1.27 (0.18-9.01) | 0.81 |
| 6-9 | 0.00% | 0.00% | - | - |
| 10-19 | 0.04% | 0.04% | 0.88 (0.20-3.94) | 0.87 |
| 20-29 | 0.04% | 0.13% | 3.15 (1.52-6.53) | <0.01 |
| 30-39 | 0.17% | 0.32% | 1.93 (1.31-2.85) | <0.01 |
| 40-49 | 0.43% | 0.90% | 2.10 (1.62-2.72) | <0.01 |
| 50-59 | 1.17% | 2.10% | 1.80 (1.50-2.16) | <0.01 |
| 60-69 | 4.44% | 5.16% | 1.16 (1.02-1.32) | 0.02 |
| 70-79 | 9.18% | 12.10% | 1.32 (1.17-1.48) | <0.01 |
| 80+ | 20.33% | 23.91% | 1.18 (1.04-1.32) | 0.01 |

**Figure 1.**
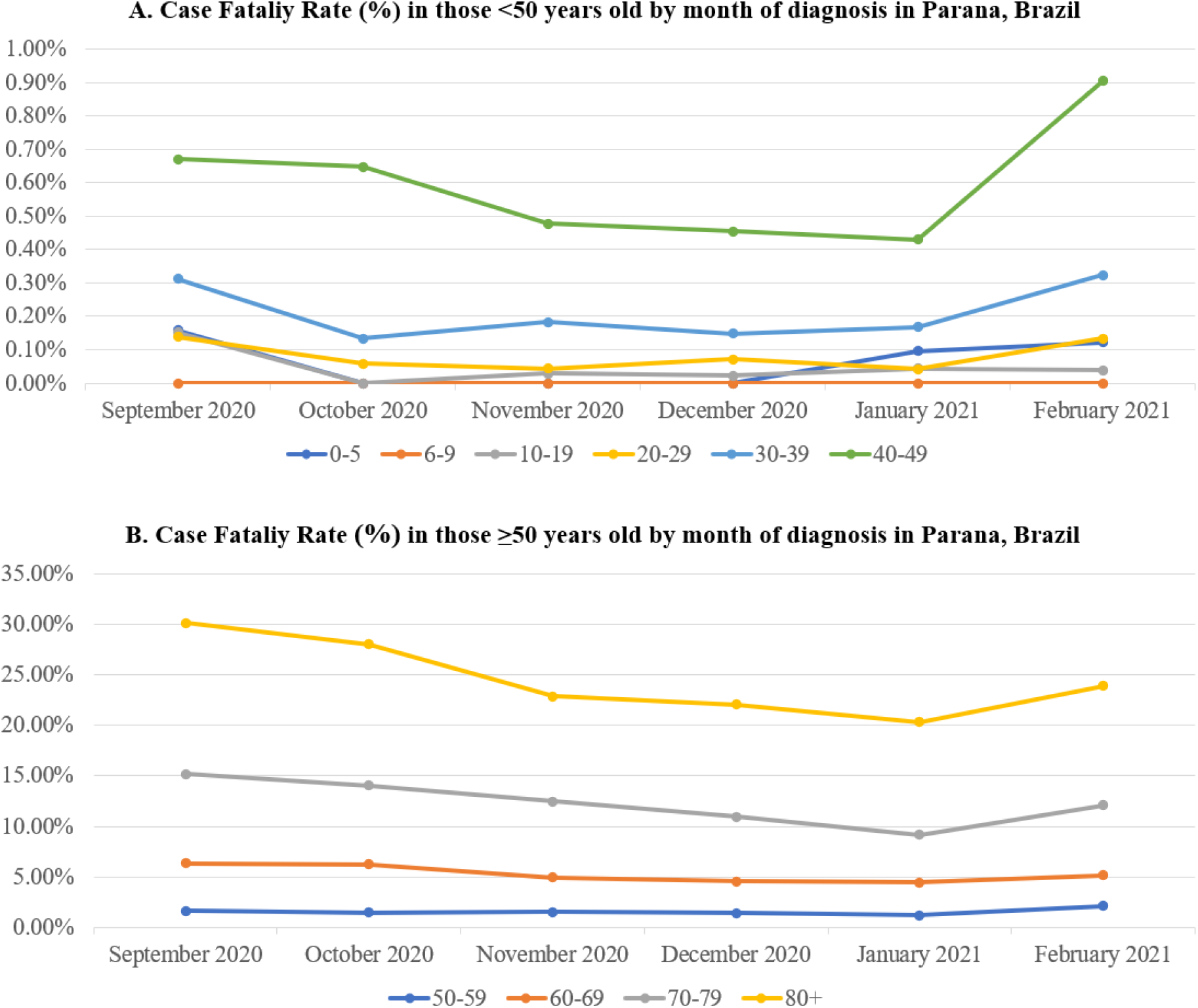
Case fatality rate in Parana, Brazil by month of diagnosis in those <50 (A) and ‘≥ 50 (B) years old.

Death RRs for each age group comparing February and January 2021 are presented in Table 1. Individuals between 20 and 29 years of age whose diagnosis was made in February 2021 had an over 3-fold higher risk of death compared to those diagnosed in January 2021. Individuals aged between 40 and 49 years also showed consistent increase in their RR, estimated at 2.10 (95% CI: 1.62-2.72), indicating those whose diagnosis was made in February 2021 had twice the risk of death of individuals diagnosed in January 2021. Similar values were observed for the 30-39 years age group, which had a RR estimated at 1.93 (95% CI: 1.31-2.85), indicating an over 90% increase in the risk of death for individuals in this age group diagnosed in February 2021 compared to the previous month.

This pattern of CFR observed over the study period was not mirrored by the monthly number of new cases, which showed expressive increases between October and December 2020, when the CFR was relatively stable, around 1.5% (Figure 2). The observed increase in CFR coincided with the second consecutive month of declining number of diagnosed cases.

**Figure 2.**
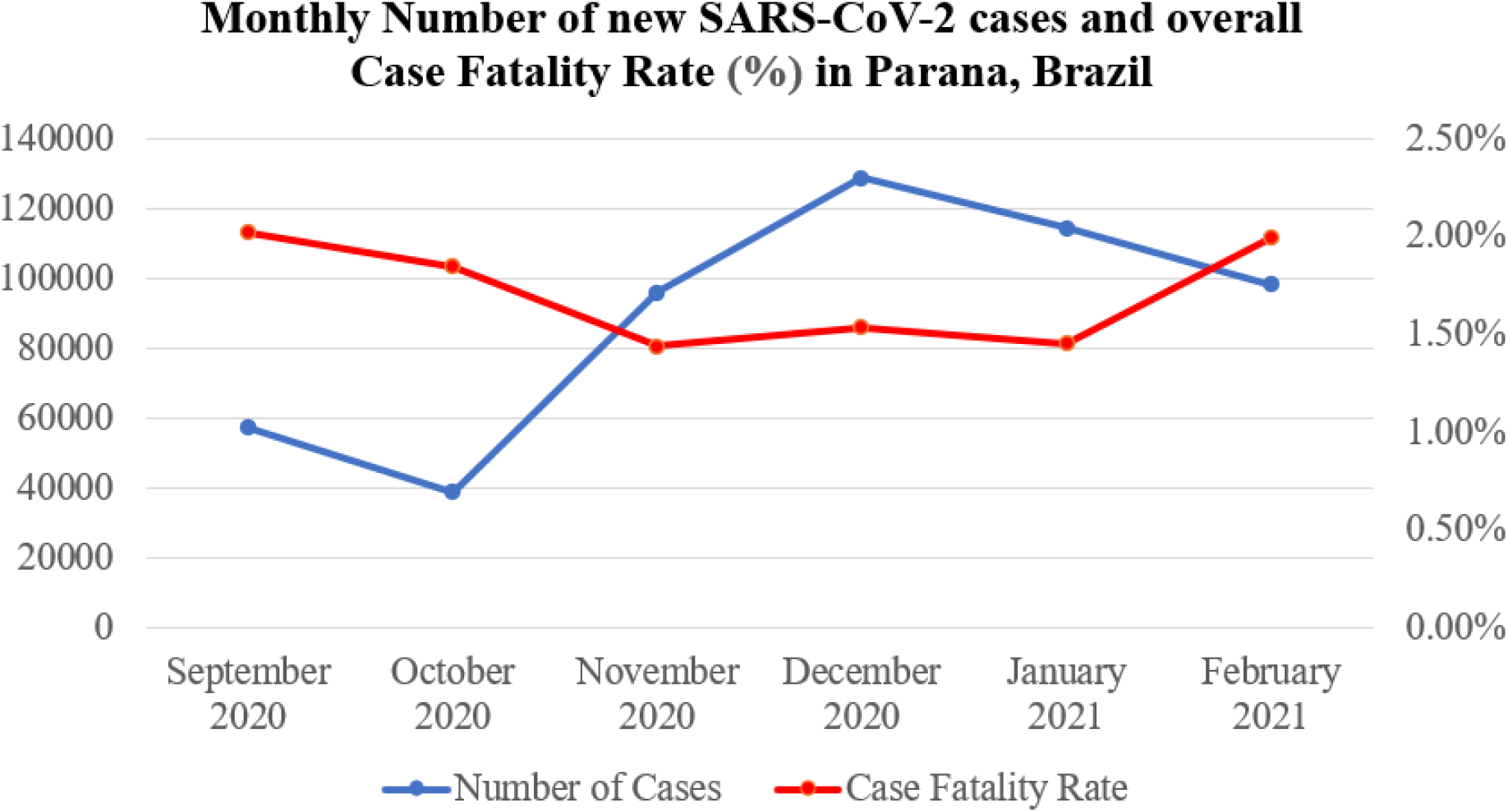
Monthly number of confirmed SARS-CoV-2 cases and fatalities in Parana, Brazil.

## DISCUSSION

In this analysis, we observed significant increases in CFR among all adult age groups in the southern state of Parana, Brazil, after identification and diffusion of the P.1 strain across the country. The most significant increases in risk of death were observed for young and middle-age adults. Importantly, no changes in CFR were observed in children or adolescents. In the state of Parana, although many regions have reached over 80% of vaccination of healthcare workers^13^, vaccines for the overall population are currently restricted to those aged 75 years of age, with administration only recently beginning,^14^ thus limiting any bias that mass vaccination may have played in the obtained results, with a minor exception for in the elderly.

The reasons for the sudden rise in CFR are not entirely clear. While in the current SARS-CoV-2 surge across Brazil an increased number of cases in younger individuals appears clearly from epidemiologic data, it is difficult to distinguish between an increased exposure/case counts in younger age groups driving overall higher raw number of fatalities at the same case fatality rate or an increased virulent form of SARS-CoV-2. Moreover, during a surge, even in well-designed clinical studies, it may be difficult to distinguish between truly increased pathogen virulence versus increased pressure on hospitals, acute care beds and other healthcare resources, thus resulting in poorer clinical outcomes as the primary driver of increased mortality rates. However, while it may be reasonable to assume that increased transmissibility of a novel strain may have resulted in overburden hospital system and healthcare resources, the rise in CFR actually coincided with a steady 2-month decline in overall cases in the state of Parana. Young and middle-aged individuals are usually less likely to take significant preventive measures and precautions, due to self-perceived lower risk of developing severe COVID-19 and onset of pandemic fatigue in recent months, thus increasing their likelihood for exposure and infection. Although this factor aspect may hence explain a sudden rise in cases among these age groups, does not thoughtfully explain a sudden increase in the CFR, especially in the presence of declining cases in this region.

While limited clinical data is available on the P.1 strain, data on variants carrying some of the same mutations, such as B.1.1.7, are associated with increased mortality.^15^ However, it is difficult to decipher from epidemiologic data alone what role a new strain may be directly playing in the observed increase in CFR. Though the P.1 strain was formally detected in only the middle of February in Parana, by March 3^rd^ it has already accounted for ∼70% of tested specimens, suggesting that circulation likely began weeks earlier than when officially detected, further clouding the epidemiological timeline.

This study was limited by the minimal scope of data available in a public health database. We choose to use date of diagnosis to associate mortality with a given month, thus it is feasible that the mortality rates, especially for February of 2021, will likely increase, as those diagnosed within that month may still be hospitalized and at risk of demise. Importantly, this study was designed only to report on recent trends in CFR among different age groups in Parana, Brazil and was not designed to assess whether or not the novel P.1 strain may be associated with increased morbidity or mortality. However, the reported CFR trends support the urgent need for well-designed clinical and epidemiologic studies, aimed to investigate potential changes in virulence or COVID-19 severity associated with the P.1 strain.

In conclusion, dramatic increases in CFR for SARS-CoV-2 infections has been noted among young and middle-aged adults in February 2021, after diffusion of P.1 across Brazil. Though these preliminary findings require immediate further investigation, they should nonetheless raise public health alarms, including the need for more aggressive local public health interventions and more rapid vaccination, and merits a global response to inhibit regional and global diffusion of this novel strain.

## Data Availability

Dataset is readily available from the website of Parana Secretary of Health.

